# Health conditions and the risk of home injury in French adults: Results from a prospective study of the MAVIE cohort

**DOI:** 10.1101/2020.10.10.20210435

**Authors:** Madelyn Yiseth Rojas Castro, Marta Avalos, Benjamin Contrand, Marion Dupuy, Catherine Sztal-Kutas, Ludivine Orriols, Emmanuel Lagarde

## Abstract

**Background:** Home injury (HI) is a significant cause of mortality and morbidity in adults of all ages. Health conditions significantly impact HI among old adults, but little is known for other adults.

**Study design:** Prospective cohort study

**Objective:** We assessed the associations between health-related factors and HI’s risk in a French study, the MAVIE cohort.

**Methods:** Poisson mixed models were fitted using health-related data information (diseases, treatments, and disabilities) at baseline and the number of injuries prospectively recorded during the follow-up, adjusting for significant socio-demographics and exposure to a range of home activities. Attributable fractions (AFs) were estimated based on RR estimations measured in the fully adjusted models.

**Results:** A total of 6,146 dwelling adults aged 15 or more were followed up for 5.1 years on average. Vertigo or dizziness (RR=2.36, 95% CI 1.06 to 5.01) and sciatica or back pain (RR=1.49, 95% CI 1.08 to 2.05) were independently associated with an increased risk of HI. These two groups of conditions showed the most significant associations among people aged 15 to 49, whereas musculoskeletal diseases other than rachialgias and arthropathies were the most significant health-related risk factor in people aged 50 and more. Sciatica or back pain represented the highest-burden of HIs in overall adults (8%) and among people aged 15 to 49 (12%).

**Conclusion:** Our results suggest that adults with musculoskeletal disorders and vertigo or dizziness symptoms have a higher risk of HI, regardless of age.

## INTRODUCTION

Home injuries (HIs) represent approximately one-third of the global burden of injuries^1 2 3^. In the EU-28, HIs caused an annual average of 85 thousand fatalities, 1.5 million hospital admissions, and 9 million emergencies between 2012 and 2014^4^. Home has been identified as the most prevalent location for injuries resulting in hospitalization and is second to roads as the location for fatal injuries^5^.

HIs affect people of all ages. However, among adults, HIs are more frequent in people aged 65 years or over. Falls are the principal mechanism of fatal and non-fatal HIs^4^. Multiple studies have identified health-related factors associated with falls in the elderly, including: lack of strength gait and balance, mobility impairment^6–8^, back pain^9 10^, arthropathies^8 11^, dizziness and vertigo^8 12 13^, obesity^14^, diabetes^7 8 11^, cardiovascular conditions^7 8 11^, chronic obstructive pulmonary disease^11^, urinary incontinence^8^, visual and hearing impairments^7^, cognition impairments caused by neurological conditions such as Parkinson’s disease, Alzheimer’s disease or other types of dementia^6–8^, and sleeping problems, anxiety or depression^6–8 11^. The presence of two or more concurrent conditions^8^ and certain medications, such as psychotropics, antidepressants, anxiolytics, sedatives or hypnotics^15^, opioids^16^, cardiac medication, diuretics, anti-diabetic medication^7^, and polypharmacy^17^ are also associated with falls in the elderly.

Among young and middle-aged adults, understanding the impact of health risk factors on HI is poor^18 19^. Regarding the risk of occupational injuries, there is some evidence of the role of hearing impairments, neurotic illness, diabetes^21^, epilepsy, and sedating medication^20^. Conditions such as back pain, sleeping problems, anxiety, depression, cardiovascular diseases, and diabetes^21^ can also be frequent in young and middle-aged adults, which justifies evaluating their impact on adults of all ages.

The purpose of this study was to assess associations of health-related factors on the risk of non-fatal HI in adults of all ages in a prospective cohort, the MAVIE cohort.

## METHODS

### Study design and recruitment

The MAVIE cohort is a web-based prospective cohort study conducted in France, with a longitudinal follow-up of *Home, Leisure, and Sports Injuries* (HLIs). All households in France and French overseas territories were eligible to participate. The recruitment process began in November 2014. The present study analyzed the data collected up to December 31, 2019. Cohort management was entirely online, including invitations, registration, and data collection. Participants were recruited through an email invitation sent to their insurees by three mutual insurance companies, press releases, social media, posters, and flyers. We asked potential participants to choose a household reference member in charge of completing a web-based questionnaire for the household. Consenting members of each household were asked to provide individual information. In an attempt to address the foreseeable underrepresentation of the elderly who may have difficulties using computers, caregivers were invited to represent and participate on behalf of one older person.

### Follow-up

Every three months, household reference members received an email reminder to report any injury event to any consenting household members during the follow-up. If no event occurred, respondents were requested to report this. An invitation to report events was also included in the monthly cohort newsletter. Still, events could be reported at any time.

### Participant data and selection

The inclusion criteria for participation in the MAVIE cohort were: i) residing in France; ii) being able to answer the questionnaires in French; iii) having access to and being able to use the Internet (at least the reference member). The baseline sample was defined as the participants aged 15 or over who answered at least one question of the individual inclusion questionnaire. We excluded those who lived in hospitals and retirement or long-term care institutions. We considered only participants who reported at least one feedback during follow-up and respondents to the daily schedule.

### Home Injuries

This study focused on unintentional home injuries or its premises. We excluded all events involving illnesses or medical symptoms. We excluded events that occurred before or on the same date as the date of consent and those for which information about the type of medical care or the circumstances of occurrence was not reported. Finally, we excluded injuries that occurred during sleeping time, considered unlikely^1^. Data included activity and location, mechanisms and type of medical care of the injury event. We defined HIs as severe when emergency care or hospital inpatient care was required.

### Individual characteristics

#### Relative time distribution

Participants were asked to report their typical daily schedule in hours for each home location. The average duration was also reported for domestic work, gardening, and do-it-yourself (DIY) activities. Time spent on each activity was categorized as *frequent* (over the 75th percentile), *occasional* or *null*.

#### Socioeconomic, demographic characteristics, and alcohol consumption

Gender, age, occupational status, education level, living situation, household incomes, and alcohol consumption were studied at baseline. Occupational status groups were *unemployed, homemakers, and retirees* and *students and employees*. Responses to education status were assigned to educational attainment levels by age groups. We considered a low level of education high school or vocational studies, or lower for participants aged 20 to 54, and primary studies or lower for other ages. Annual household incomes were assigned to classes according to the French population’s percentiles reported in 2015 (Low: ≤ 30th percentile, Middle: 30th-80th percentile, High: ≥ 80th percentile). The categories of the frequency of alcohol consumption were less than two times a week, and two or more times a week.

#### Health conditions and medications

At inclusion, participants were invited to report their history of diseases and treatments over the previous 12 months.

Medical conditions were grouped into the following clinically homogeneous categories: cardiovascular diseases, respiratory diseases, digestive diseases, genitourinary diseases, endocrine diseases, sciatica or back pain, arthropathies, other conditions of the musculoskeletal system, eye diseases, depression, anxiety, sleeping disorders (including treatment with antidepressants, anxiolytics or hypnotics), migraine or headache, vertigo or dizziness, chronic mental diseases (Parkinson’s disease, Alzheimer’s disease or other senile dementia, epilepsy, and antiepileptic medicines), and cancer. Participants were also asked to report whether they were suffering from impairments or disabilities related to vision, hearing, mental, intellectual, or psychological, and mobility.

We defined polypharmacy as a reported number of medications of 4 or more^17^, and comorbidity as the report of two or more diseases^22^ at any time in the previous 12 months. The history of injuries (excluding sports injuries) over the past 12 months was also reported.

## Statistical analyses

Data were analyzed using R version 3.6.1 (The R Foundation for Statistical Computing, Vienna, Austria)

We defined the follow-up period (FP) from the consent date to the latest date among the following: the latest news statement date, the questionnaires update date, the cohort exit date, the death date. We calculated the proportion of time spent awake at home (PTAH) as the proportion of hours on a weekday or weekend day when the person was neither away from home nor sleeping. We considered the *time-at-risk* as:

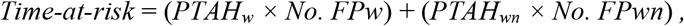

where *PTAH*_*w*_ was the time spent awake at home regular weekday, *PTAH*_*wn*_ on a regular weekend day, *No. FP*_*w*_ the number of weekdays in the follow-up period and *No. FP*_*wn*_ the number of weekend days. We calculated the proportion of *time-at-risk* as:

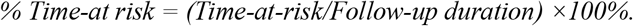

### Risk factor analysis

Poisson mixed models were used to fit the number of HIs during the estimated time at risk. Random effects were included to take into account the cluster structure of data in households. All models were fitted using the Template Model Builder R package^23^. Relative risks (RRs) were calculated conditionally upon the random effects, and the 95% confidence intervals were calculated using the profile method^24^. We adopted a complete case approach.

We fitted individual crude regression models to each health condition. Then, we conducted a two-step variable selection procedure. First, we adjusted each model by age, gender, and history of injuries in all models, and we selected the most important associations (*p*_*adj*_ < 0.10). Second, we fitted the models to each health-related factor, adjusting for significant adjustment variables selected in the previous step. Finally, we selected the most important associations for the fully adjusted model (excluding comorbidity). Benjamini-Hochberg corrected p-values were computed ^25^. The same methodology was used to construct models stratified by two distinct age groups (15 to 49 years old and 50 years old or more).

The attributable fraction (AF) was estimated using the adjusted RR of the fully adjusted models, as suggested by Flegal and colleagues, 2006^26^.

## RESULTS

Between November 2014 and December 2019, 11,420 adults registered in the MAVIE cohort and responded to baseline questionnaires. Among them, 9,429 were household reference members representing themselves and/or another household member. Sixty-six participants reported that living in a retirement institution, and 3,524 participants did not provide follow-up data. Another 1,684 participants did not provide their daily schedule (Figure 1). The sample size was 6,146 participants from 5,122 households.

**Figure 1.**
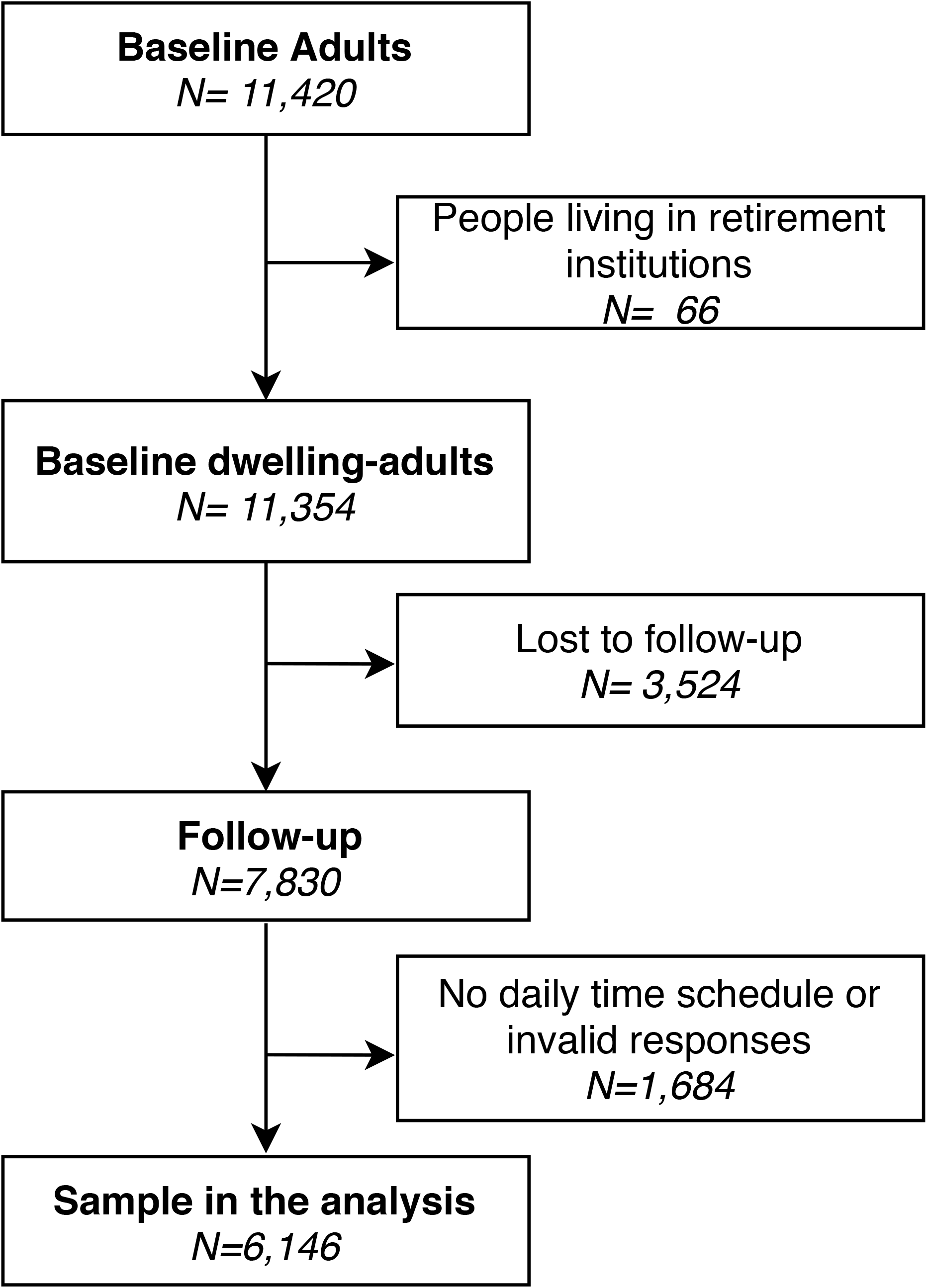
Flowchart of study participants

The median follow-up was 4.0 years (Q1=3.6, Q3=4.5). The overall follow-up duration was 4,800 persons-time. The loss to follow-up was 13%.

Among the 6,146 individuals of the study, 244 left the cohort during the follow-up. Nineteen of them were reported by the reference member to have died, 16 from illness, and three of unknown causes. Thirty-one participants left the study due to changes in household composition. The remaining 191 participants did not report any reasons for leaving the study. Socioeconomic, demographic characteristics, and alcohol consumption levels were similar to those of the study sample and baseline sample (Table 1). However, dropout was more frequent among young adults, students, and employees, and among members of low-income households.

**Table 1.**
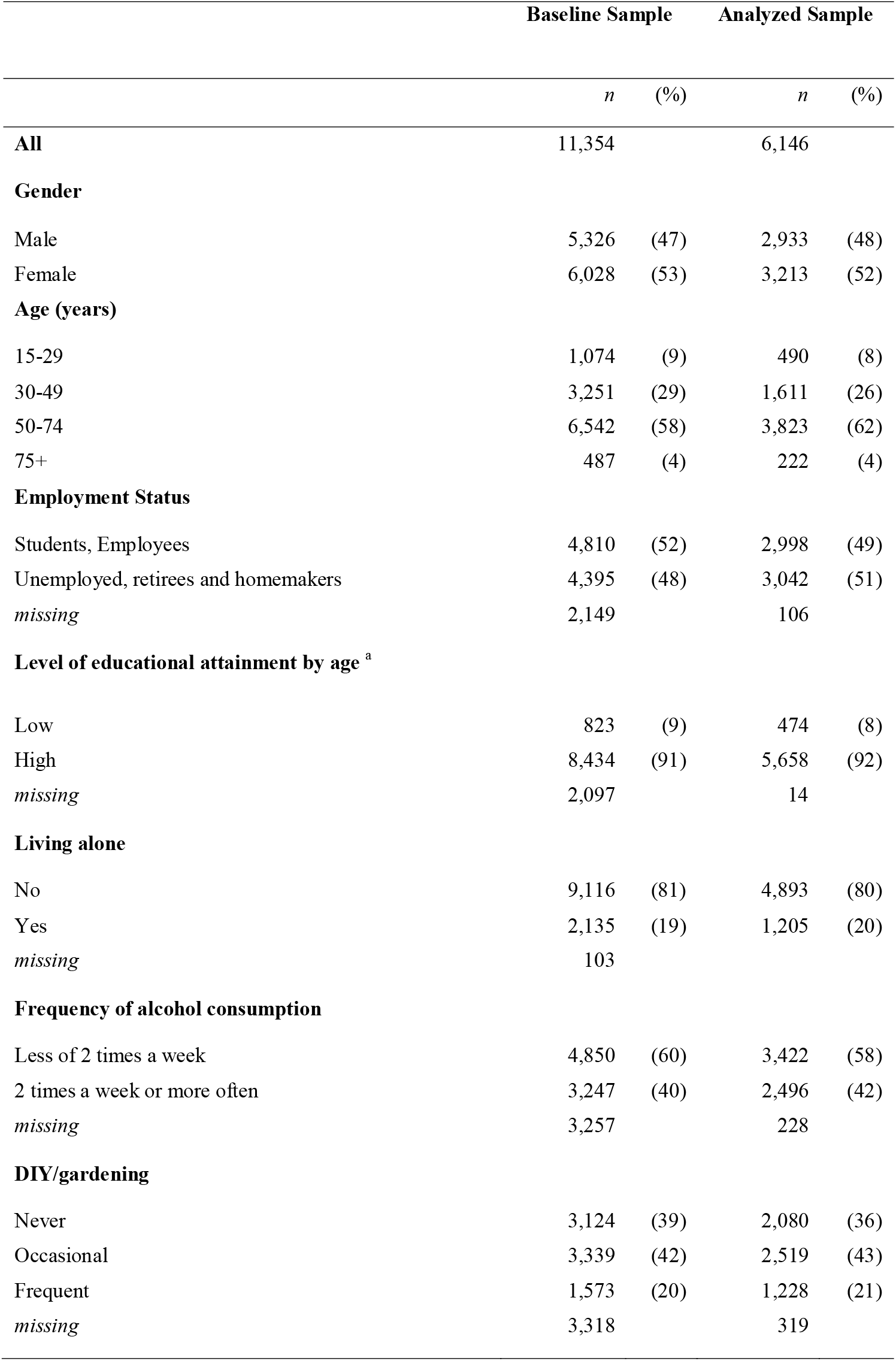
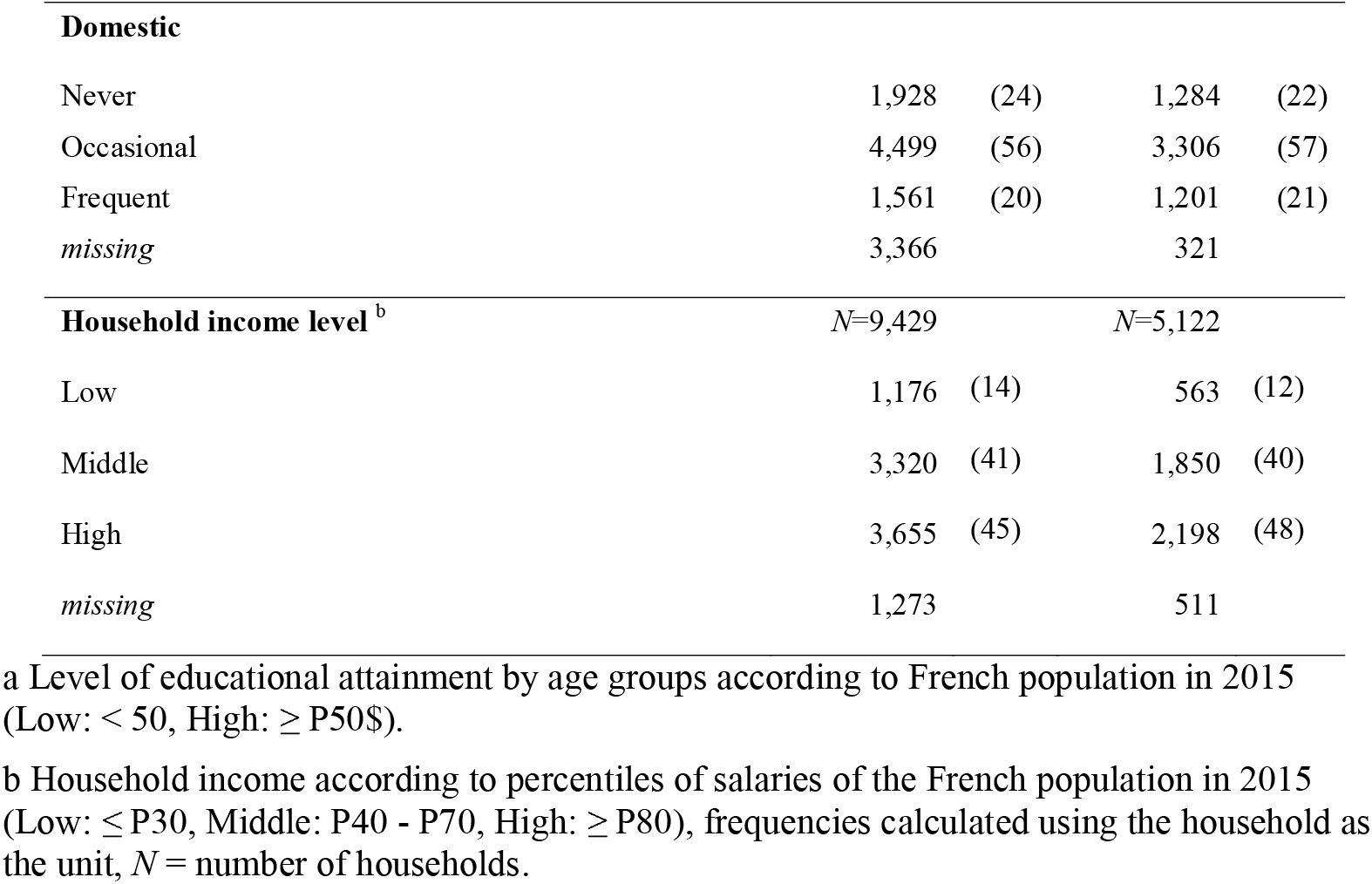
Description of the demographic, socioeconomic and other characteristics at inclusion time of MAVIE dwelling-adults: baseline and analyzed sample

### Baseline characteristics

Socioeconomic, demographic characteristics, and alcohol consumption Table 1 shows the socioeconomic, demographic characteristics, and alcohol consumption of the sample at baseline. Sample characteristics differed from those of the overall French population (INSEE Census, 2015). Participants aged 50 to 74 were overrepresented (62% vs. 35% in the French population, 2015). Students and employees were underrepresented (49% vs. 57%). People with low education attainment level were underrepresented (8% vs. 44%). Finally, only 12% of household reference members reported an annual income lower than the 30th percentile of French households.

The median proportion of time spent at home awake was 48% (Q1=36%, Q3=58%). Adults older than 50, unemployed, retirees, homemakers, and those who reported gardening, DIY, and domestic activities were those who spent the most time at home (Supplementary material 1).

### Health conditions, diseases, and medications

Among the 6,146 participants, 67% reported at least one health condition (Figure 2). They were older, with a median age of 60 compared to 35 for those who did not report any health condition. Figure 2 shows the most frequently reported health conditions by age group. Almost half of the participants (48%) reported more than one health condition. The most frequently reported conditions among adults aged 15 to 49 years were depression, anxiety, or sleeping disorders, and among adults aged 50 and more, cardiovascular diseases. Eight percent of participants reported at least one type of injury in the last 12 months.

**Figure 2.**
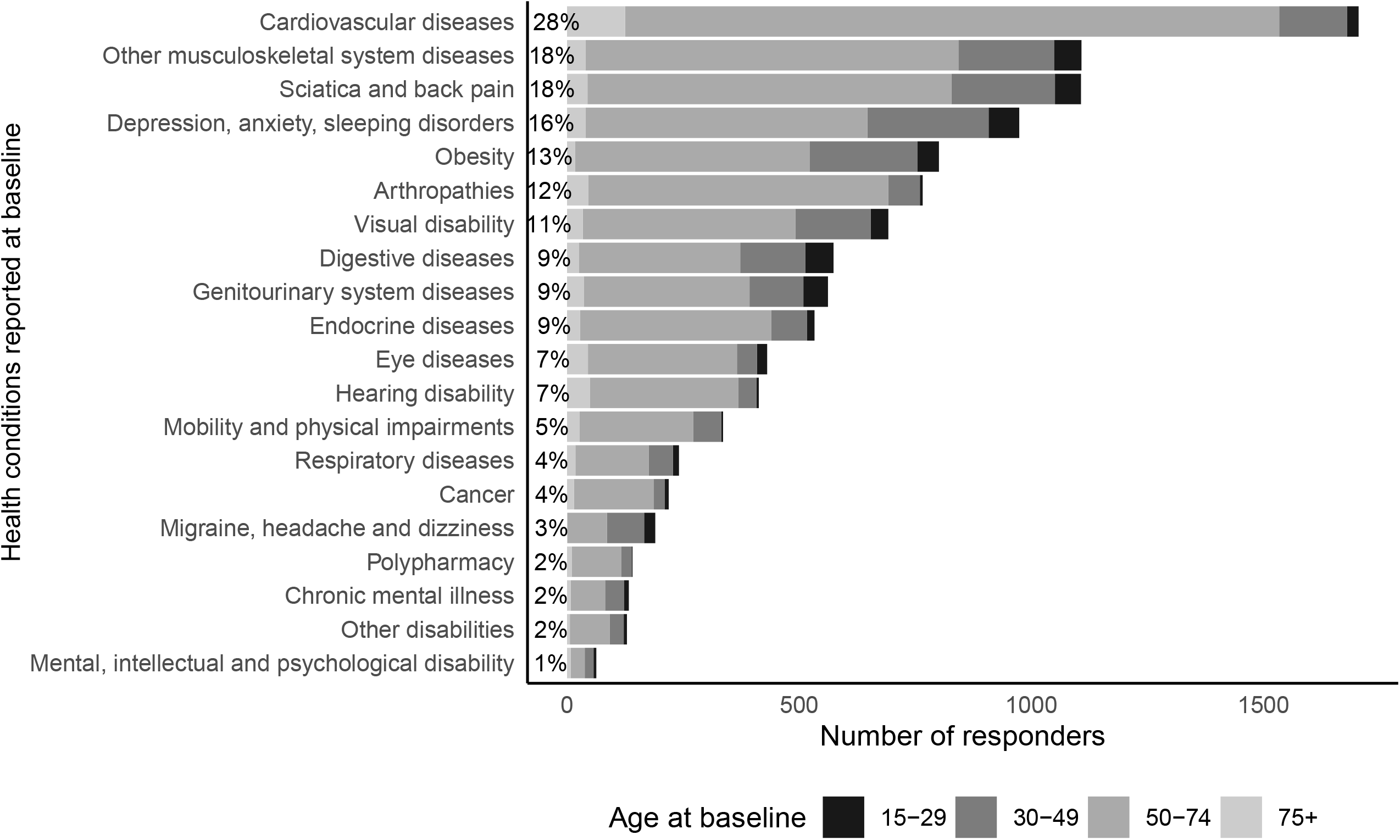
Self-reported health conditions at inclusion: Diseases reported during the 12 months prior to baseline, disabilities, and obesity at the time of inclusion by the analyzed dwelling-adults of the MAVIE cohort, stratified by age groups (at inclusion). Sample size N = 6,146

### Home Injuries

During follow-up, 12% of the participants reported at least one HI, of which 946 occurred while awake. Most of HIs were minor injuries, and only 28% required either hospitalization or emergency care. Falling was the main mechanism of HI (39 %), domestic and gardening activities (42%), the most frequents activities, and the garden (25%) the most frequent location (Supplementary Material 2).

### Health risk factor analysis

Adjusting for the selected variables of block 1 (Table 2), vertigo or dizziness, sciatica or back pain, arthropathies, other musculoskeletal system diseases, depression, anxiety or sleeping disorders, and comorbidity were associated with an increased risk of HI in adults of all ages with a small or moderate effect (Block 2, Table 2). In the fully adjusted model (Table 3), vertigo or dizziness and sciatica or back pain remained associated with an increased risk of HI and with an AF of 1.9% and 8.1%, respectively. The contribution of depression, anxiety, and sleeping disorders was also important (AF=5.5), but the association was weaker than the conditions named above (RR = 1.28, 95% CI 0.96 to 1.70).

**Table 2.**
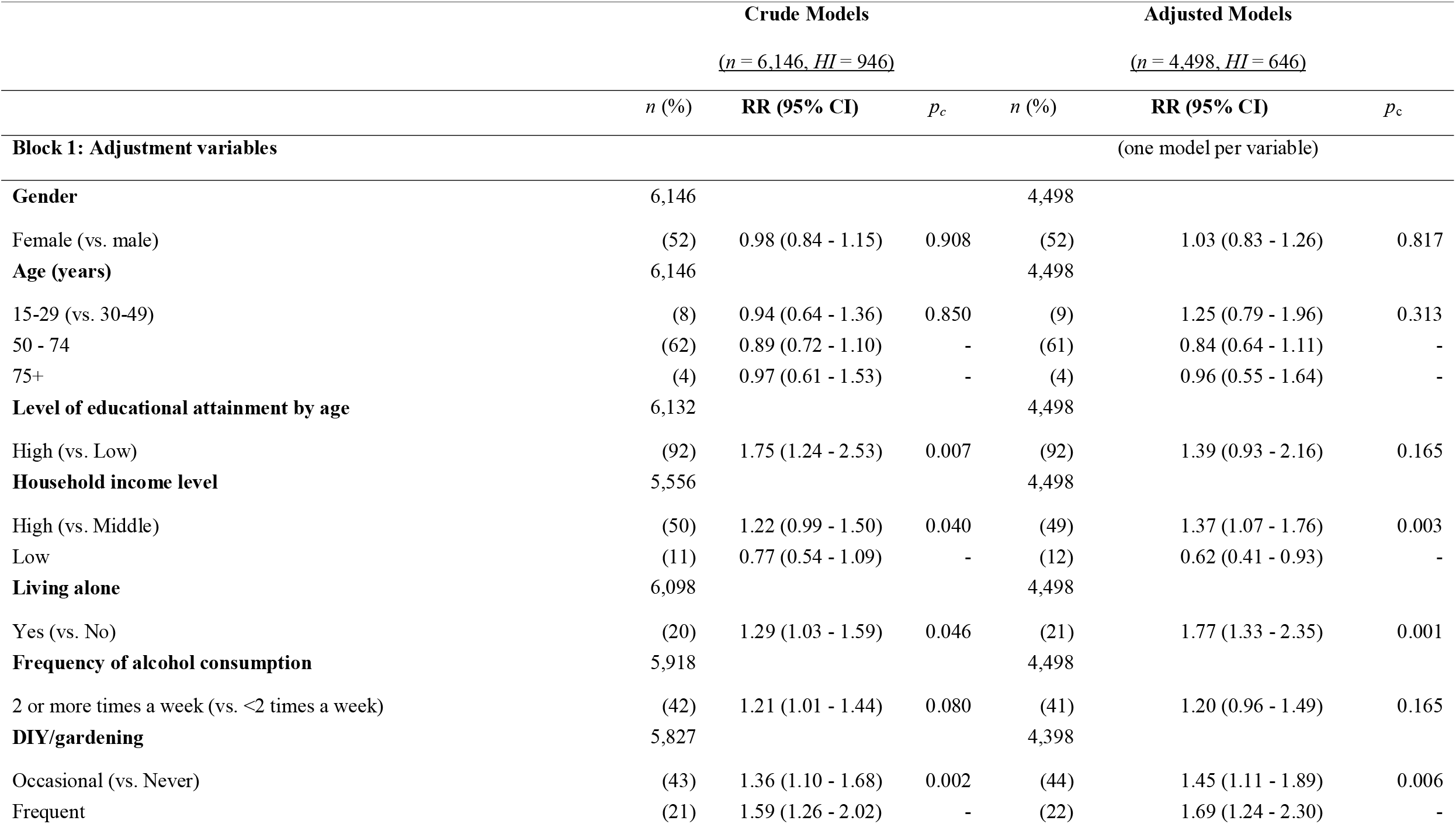
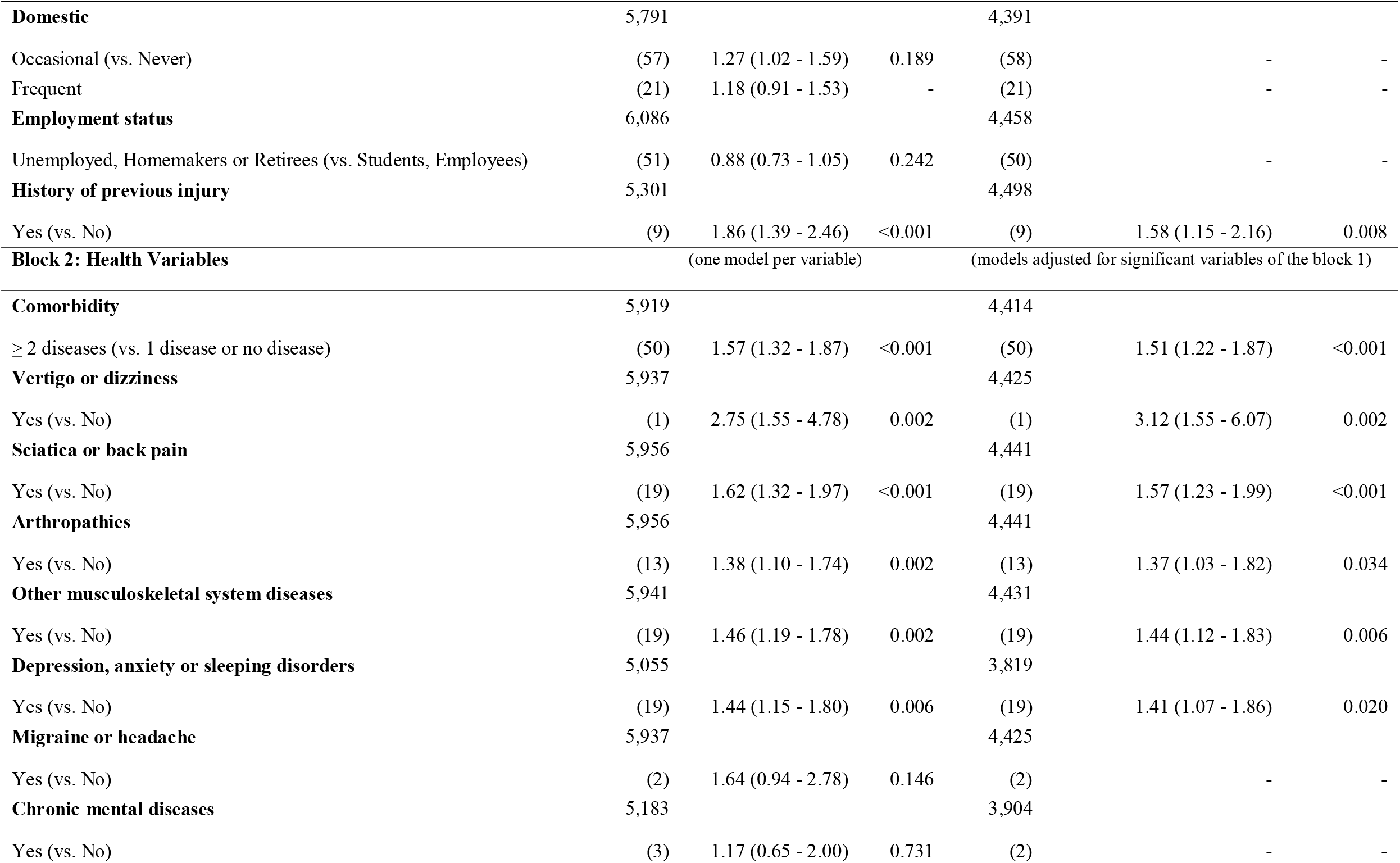
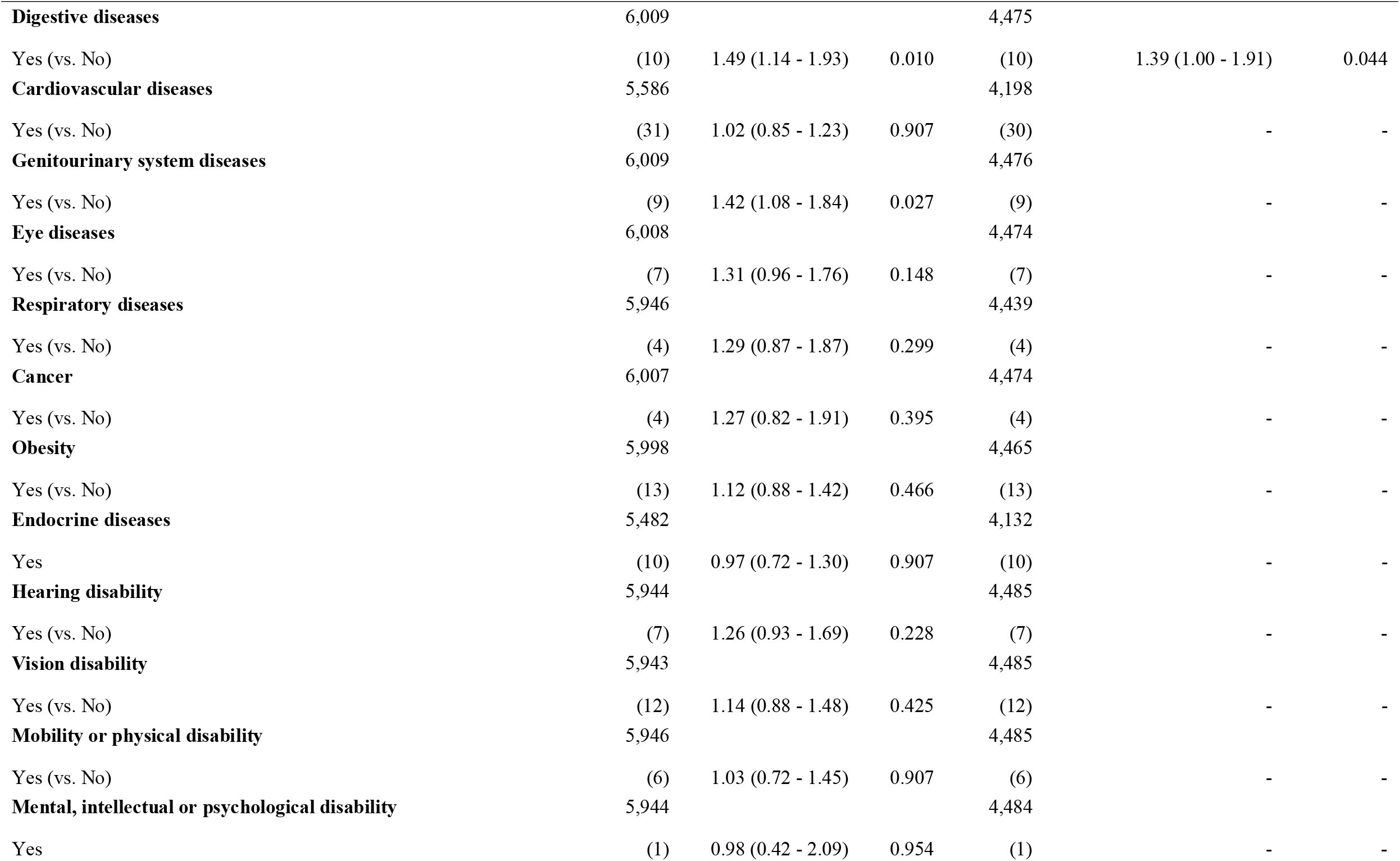
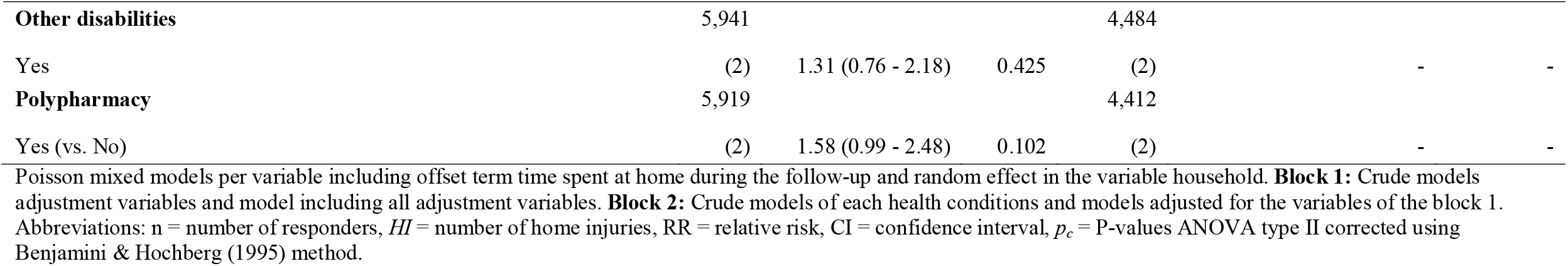
Individual factors associated with the incidence of HI, relative to the time at risk at home, in adults of the MAVIE cohort

**Table 3.**
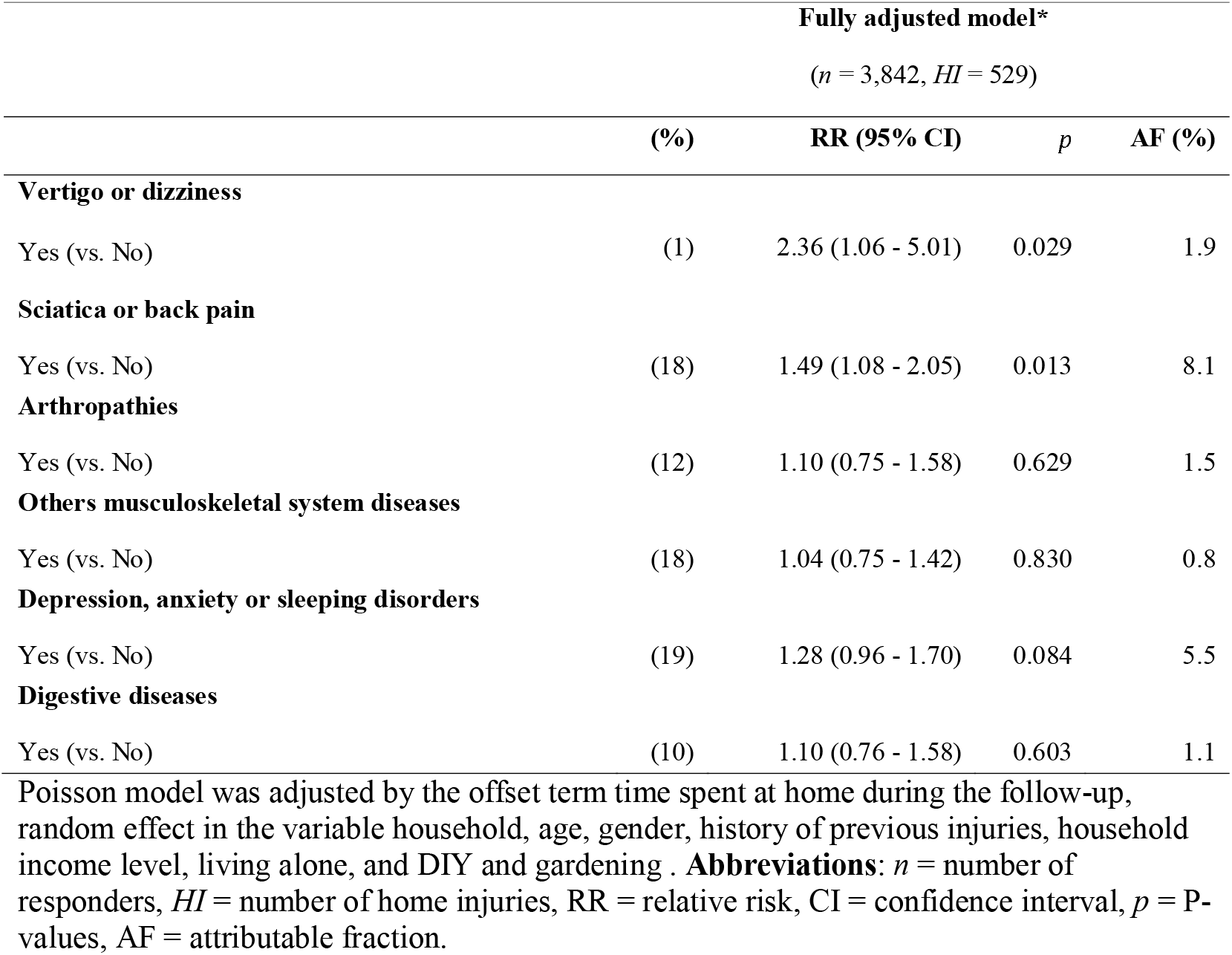
Factors associated with the incidence of HI, relative to the time at risk at home, in adults of the MAVIE cohort *(fully adjusted model)*

Results of the fully adjusted models were similar for participants aged 15 to 49, but the effects of sciatica or back pain and vertigo or dizziness effects were higher than among adults of all ages (Table 4). The category of other musculoskeletal system diseases covered the only conditions significantly associated with the risk of HI among participants aged 50 or more (Table 4). All models converged, and their residuals were validated, discounting problems, such as over/underdispersion and zero inflation.

**Table 4.** Factors associated with the incidence of HI, relative to the time at risk at home, in adults of the MAVIE cohort by age group

| Adjusted models |  |  |  | Fully adjusted models |  |  |  |
| --- | --- | --- | --- | --- | --- | --- | --- |
| | (%) | RR (95% CI) | $p_c$ | (%) | RR (95% CI) | $p$ | AF (%) |
| **Age 15-49** | *(n = 1,887, HI = 208)* |  |  | *(n = 1,804, HI = 205)* |  |  |  |
| **Sciatica or back pain** |  |  |  |  |  |  |  |
| Yes (vs. No) | (14) | 2.40 (1.49 - 3.91) | <0.001 | (14) | 2.26 (1.40 - 3.65) | 0.001 | 11.9 |
| **Vertigo or dizziness** |  |  |  |  |  |  |  |
| Yes (vs. No) | (2) | 4.95 (1.76 - 13.75) | 0.002 | (1) | 4.59 (1.48 - 14.13) | 0.007 | 3.2 |
| **Comorbidity** |  |  |  |  |  |  |  |
| ≥2 diseases (vs. 1 disease or not disease) | (41) | 2.15 (1.45 - 3.22) | <0.001 | - | - | - | - |
| **Age 50+** | *(n = 2,915, HI = 472)* |  |  | *(n = 2,59, HI = 468)* |  |  |  |
| **Others musculoskeletal system diseases** |  |  |  |  |  |  |  |
| Yes (vs. No) | (21) | 1.57 (1.19 - 2.07) | 0.005 | (22) | 1.45 (1.05 - 1.98) | 0.023 | 9.3 |
| **Sciatica or back pain** |  |  |  |  |  |  |  |
| Yes (vs. No) | (21) | 1.36 (1.03 - 1.80) | 0.031 | (21) | 1.09 (0.78 - 1.52) | 0.602 | 2.2 |
| **Arthropathies** |  |  |  |  |  |  |  |
| Yes (vs. No) | (17) | 1.37 (1.02 - 1.84) | 0.036 | (18) | 1.14 (0.81 - 1.59) | 0.458 | 2.7 |
| **Comorbidity** |  |  |  |  |  |  |  |
| ≥ 2 diseases (vs. 1 disease or no disease) | (53) | 1.35 (1.05 - 1.75) | 0.036 | - | - | - | - |

Poisson mixed models including offset term time spent at home during the follow-up and random effect in the variable household. Adjustment variables: **Age 15-49** (gender, history of injuries), **Age 50+** (gender, history of injuries, living alone, household income level, DIY/gardening, frequency of alcohol consumption). Abbreviations: *n* = number of responders, *HI* = number of home injuries, RR = relative risk, CI = confidence interval, AF = attributable fraction, $p_c$ = P-values ANOVA type II corrected using Benjamini & Hochberg (1995) method, $p$ = P-values

## DISCUSSION

To our knowledge, we present here the first study to evaluate the association of health conditions with the risk of non-fatal HIs in adults of all ages. We followed up 6,146 cohort participants residing in French households, of whom 12% suffered at least one HI over an average of 5.1 years. The same proportion was 14% among those who reported at least one health condition. Among the 21 health conditions reported at inclusion, we identified sciatica or back pain problems and vertigo or dizziness as the main risk factors, with an estimated AF of 8.1% and 1.9%, respectively.

This unprecedented finding of substantially increased risk of HI among adults under 50 years old with sciatica or back pain (RR=2.26) and vertigo or dizziness (RR=4.59) is striking because of these high association measures, but also because of a substantial estimated AF (11.9% and 3.2%, respectively). For people over 50 years old, musculoskeletal diseases (other than rachialgia and arthropathies) are the only condition that remained significant in the fully adjusted model. These same factors associated with mobility emerged when we analyzed falls and shocks only, whereas this was not the case for other HIs. That suggests that most health conditions affecting the risk of HI are increasing the risk of falling. Other factors usually associated with injuries and falls, such as gender, age, vision and hearing impairments, polypharmacy, and alcohol consumption, did not appear to be associated with an increased risk of HI in this study.

Musculoskeletal conditions (including back pain) and sciatica are well-known risk factors for occupational injury, likely due to overexertion. Overexertion was reported only in the 18% of injuries of people suffering from musculoskeletal conditions. Consistent with previous findings, the effects of arthropathies (RR=1.37) and other musculoskeletal conditions (RR=1.57) were more relevant among adults over 50 years old^8 11^. In contrast to previous studies, back pain did not appear to be associated with an increased risk of HI in adults over 50^9,10^. One possible explanation is that older people may favor chronic conditions over problems such as back pain when self-reporting. We hypothesize that poor posture might affect housework performance, increasing general injury susceptibility. Decreased cognitive function due to chronic pain^27^ or opioid use^16^ might also raise HI risk.

Vertigo or dizziness increased the risk of HI among young adults, but not among those aged 50 or more. This group of symptoms may be related to vestibular migraine, benign paroxysmal positional vertigo, and Meniere’s disease^28^, rather than age-related conditions such as Parkinson’s disease, or cerebellum and oculomotor brain stem syndrome^29^. Other possible causes of vertigo, such as hypoglycemic episodes or postural hypotension, have been partially controlled for by adjusting for other conditions such as endocrine diseases and cardiovascular diseases.

We found a slightly increased risk of HI among those reporting depression, anxiety, or sleeping disorders. Consistently, Palmer and colleagues (2018) highlighted an increased risk of occupational injury among people with emotional conditions, and a moderate effect among sedatives consumers^20^. Another explanation might be the difficulties in assessing risks and a reduced motivation to correct them in people who have affective psychological disorders. Such an effect was observed for patients with Parkinson’s disease^30^.

Unlike other studies^20 31^, we found no association regarding age and disability. Because we could account for the time spent at home, our results confirm that exposure time is the main explanation^1 32–34^. A relevant self-perception of disability allows compensation for an increased risk of injury, enabling enhanced care in daily activities and avoiding risky tasks and domestic hazards. People with moderate conditions, more active in domestic work, and less aware of risks might be more exposed to hazards together they might insufficiently regulate their behavior. On the other hand, these effects may be mitigated by the beneficial impacts of domestic activities^35^, for example, by improving executive functions^36^.

Lessons for prevention from our results include better awareness of risks at home for people with diseases related to the musculoskeletal system. Our results also highlight the preventive role of physicians when dealing with vulnerable people. However, HI risk related to the level of activities should not discourage leisure activities and help raise awareness and adaptation of the living environment to the risks by mapping foreseeable hazards.

### Limitations and Strengths

Common limitations observed in volunteer-based cohorts and e-cohorts are low response rates, volunteer bias, loss of follow-up, and self-administered questionnaires, leading to selection biases and missing and selective answers. We made continuous efforts to address representativeness, loss to follow-up, and quality of information issues. However, young adults and people from low socioeconomic groups and levels of educational attainment were underrepresented. An overestimation of the prevalence and impact of some diseases is probable. The difficulty of diagnosing the conditions, the ability to recall, the willingness to report (mostly when communicated by reference members), physical and mental quality of life may also affect the report^37^. Besides, this study design does not allow us to disentangle the effects of health conditions and treatments. Moreover, information about disease severity, risk behaviors, and risk awareness was not available. Finally, we measured health conditions at a single time point; health changes were not assessed over time.

## Conclusion

Despite these limitations, the MAVIE cohort is the injury observatory with the most detailed sociodemographic, contextual, and exposure information to study risk factors of HLIs in France. This information enabled us to control for potential confounders and account for exposure time, the latter being a rare opportunity.

## Data Availability

Reasonable requests for patient level data should be made to the corresponding author and will be considered by INSERM and Calyxis responsible for any decision regarding the possible reuse of the data.

## FOOTNOTES

## Acknowledgements

The authors express their thanks to all IETO team members for their help, especially Juan Naredo Turrado, Jeanne Duchesne, Li Lu and Marie-Odile Coste.

## Contributors

All authors contributed to the study conception and design. Methodology: MR, MA and EL. Formal analysis: MR, Investigation: MR, MA, LO and EL. Writing original draft: MR Writing review & editing: MA, BC, MD, CS, LO and EL. Supervision: EL, MA. Project administration: EL, CS and MD. Funding acquisition: EL, CS, and MD.

## Ethical approval

The French Data Protection Authority approved the protocol of this study. The study is declared to the CNIL under file number 912292. Identifying data were stored on servers located in a different location from those hosting the main database. Electronic informed consent was collected from all adult participants. participation of children was done under the responsibility and with the consent of a legal guardian.

## Data Sharing

Reasonable requests for patient level data should be made to the corresponding author and will be considered by. INSERM and Calyxis, *Pôle d’expertise du risque* responsible for any decision regarding the possible reuse of the data.

## Patient and Public Involvement

Since inclusion, participants to the MAVIE cohort have received summaries of the main conclusions obtained and have at their disposal an application the MAVIE Lab that allows them to experiment with the results.

## Transparency

EL (corresponding author) affirm that the manuscript is an honest, accurate and transparent account of the study being reported; that no important aspects of the study have been omitted; and that any discrepancies from the study as originally planned (and, if relevant, registered) have been explained.

## What is already known on the subject?

- There is evidence that a large number of health conditions increase home injury risk in older adults.
- Musculoskeletal system diseases increase the risk of falls at home in older adults through weakness, loss of posture and balance.
- There is a lack of evidence on how health conditions affect the risk of home injury in younger adults.

## What this study adds?

- Our results suggest a moderate to high effect of musculoskeletal system diseases and vertigo or dizziness symptoms over home injury risk regardless of age.
- Back pain or sciatica is the group of conditions with the highest attributable fraction of home injury risk among all adults and adults under 50 years old.

