## Supplementary material for "Health conditions and the risk of home injury in French adults: Results from a prospective study of the MAVIE cohort"

**Supplemental material** 1–Percentage of time-at-risk and incidence rate of home injuries by the time at home, in adults of the MAVIE cohort, by demographic, socioeconomic and others characteristics

|  | **% Time-at-risk ^a^**  **Q50 (Q25 – Q75)** | **HI** | **IR (95% CI) ^b^** |
| --- | --- | --- | --- |
| **All** | 48 (36 - 58) | 946 | 81 (76 - 87) |
| **Gender** |  |  |  |
| Male | 49 (36 - 59) | 460 | 80 (73 - 88) |
| Female | 46 (35 - 57) | 486 | 83 (76 - 90) |
| **Age (years)** |  |  |  |
| 15 - 29 | 36 (30 - 43) | 60 | 89 (68 - 115) |
| 30 - 49 | 36 (32 - 44) | 222 | 89 (78 - 102) |
| 50 - 74 | 53 (43 - 60) | 620 | 78 (72 - 84) |
| 75+ | 59 (53 - 63) | 44 | 87 (64 - 117) |
| **Employment status** |  |  |  |
| Students, Employees | 36 (31 - 43) | 403 | 89 (80 - 98) |
| Unemployed, retirees and homemakers | 56 (50 - 62) | 527 | 76 (69 - 82) |
| Level of educational attainment by age ${}^{c}$^c^ |  |  |  |
| High | 47 (35 - 58) | 896 | 84 (78 - 89) |
| Low | 50 (37 - 60) | 49 | 54 (40 - 72) |
| Living along |  |  |  |
| No | 48 (36 - 58) | 716 | 77 (71 - 83) |
| Yes | 47 (35 - 57) | 222 | 99 (86 - 113) |
| **Frequency of alcohol consumption** |  |  |  |
| Less of 2 times a week | 44 (35 - 57) | 479 | 77 (70 - 84) |
| 2 times a week or more often | 51 (37 - 59) | 448 | 89 (81 - 98) |
| **DIY/gardening** |  |  |  |
| Never | 42 (33 - 55) | 227 | 63 (55 - 72) |
| Occasional | 46 (35 - 58) | 403 | 84 (76 - 93) |
| Frequent | 55 (45 - 61) | 258 | 97 (85 - 109) |
| **Domestic** |  |  |  |
| Never | 52 (38 - 60) | 172 | 66 (57 - 77) |
| Occasional | 43 (34 - 56) | 511 | 86 (79 - 94) |
| Frequent | 52 (40 - 59) | 200 | 83 (72 - 95) |
| **Household income level ^d^** |  |  |  |
| High | 46 (35 - 58) | 462 | 89 (81 - 97) |
| Middle | 48 (36 - 58) | 319 | 76 (68 - 85) |
| Low | 48 (37 - 58) | 68 | 60 (46 - 76) |

a Percentage estimated of time at home “awake” during the follow-up time.

b Injury incidence rate per 1.000 person-year spent at house awake.

c Level of educational attainment by age groups in 2015 Low : <P50, High: ≥ P50).

d Household income according to percentiles of salaries of the French population in 2015 (Low: ≤ P30, Middle: P40 − P70, High: ≥ P80).*Abbreviations:* HI = number of home injuries, IR incidence rate, CI confidence interval. Sample size N=6,146

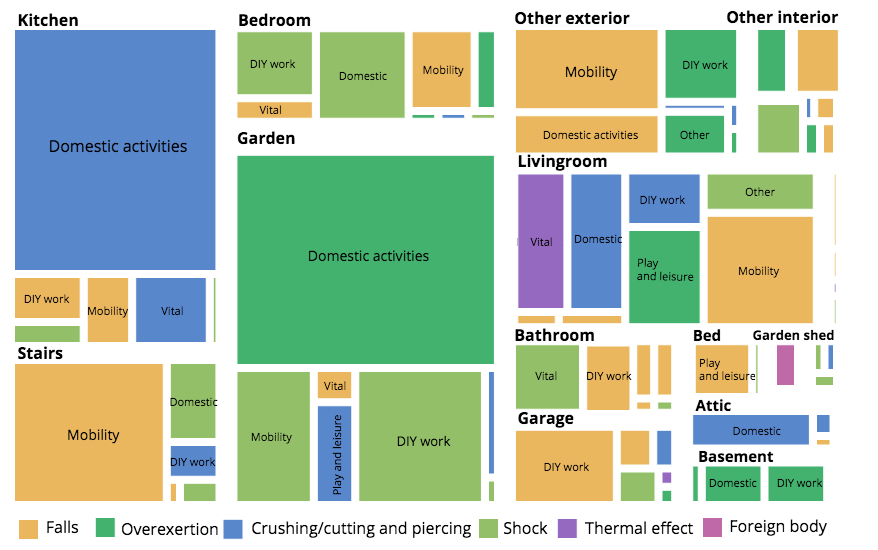

**Supplemental material** 2 *–* Activities, places and mechanisms of injury at home. Mosaic graph, surface areas are proportional to the number of injuries events reported by activity and location and the colors represents main mechanism of the injury.
